# The Student Patient Alliance: Development and formative evaluation of an initiative to support collaborations between patient and public involvement contributors and doctoral students

**DOI:** 10.1101/2023.01.26.23285050

**Authors:** Gwenda Simons, Rebecca Birch, Joanne Stocks, Elspeth Insch, Rob Rijckborst, Georgiana Neag, Heidi McColm, Leigh Romaniuk, Claire Wright, Bethan E Phillips, Simon W Jones, Arthur G Pratt, Stefan Siebert, Karim Raza, Marie Falahee

**Author notes:** **CORRESPONDING AUTHOR:** Dr Marie Falahee, Rheumatology Research Group, Institute of Inflammation and Ageing College of Medical and Dental Sciences, University of Birmingham Research Laboratories, Queen Elizabeth Hospital, Birmingham B15 2WB, United Kingdom.

## Abstract

**Background:** While the integration of patient and public involvement (PPI) in clinical research is now widespread and recommended as standard practice, meaningful PPI in pre-clinical, discovery science research is more difficult to achieve. One potential way to address this is by integrating PPI into the doctoral training programmes of discovery science postgraduate students. This paper describes the development and formative evaluation of the Student Patient Alliance (SPA), a programme developed at the University of Birmingham that partners PPI contributors with doctoral students.

**Methods:** Following a successful pilot of the SPA by the Rheumatology Research Group at the University of Birmingham, the scheme was implemented across collaborating Versus Arthritis / MRC centres of excellence at a number of different collaborating centres. Students were partnered with PPI contributors, provided with initial information and guidance, and then encouraged to work together on research and public engagement activities. After six months, students, their PPI partners and the PPI coordinators at each centre completed brief surveys about their participation in the SPA.

**Results:** Both students and their PPI partners felt that taking part in SPA had a very positive impact. Students reported an increased understanding of PPI and patient priorities and reported improved public engagement and communication skills. Their PPI partners reported a positive impact of the collaboration with the students. They enjoyed learning about the student’s research and contributing to the students ‘personal development. PPI coordinators also highlighted the benefits of the SPA, but noted some challenges they had experienced, such as matching students with PPI partners.

**Conclusions:** The SPA was valued by students and PPI partners, and it is likely that initiatives of this kind would enhance students’ PPI and public engagement skills and awareness of patients’ experiences on a wider scale. However, appropriate resources are needed at an institutional level to support the implementation of effective programmes of this kind on a larger scale.

## BACKGROUND

The involvement of members of the public or patients in research is often referred to as Patient and Public Involvement (PPI) and can be defined as “*research carried out ‘with’ or ‘by’ members of the public rather than ‘to’, ‘about’ or ‘for’ them”* (1). PPI is widely recognised to enhance research relevance, accountability, acceptability, quality, and dissemination (including to lay audiences), whilst also increasing the chances of the research being implemented (2, 3). Not only can PPI be beneficial for research, but it can, and should be beneficial to the PPI contributors themselves, and the people they represent, giving them a voice that matters (3) and in many cases the feeling that they can ‘give back’ to the community. It is also important to note that for many research funders, PPI has become a key requirement (4).

The rheumatology research community has played a pioneering role during the development of PPI in (clinical) research and has provided an exemplar for many other specialities in implementing and developing PPI strategies (5). For example, the European Alliance of Associations for Rheumatology (EULAR) developed recommendations (6) and resources (7) to support PPI in rheumatology research, with other rheumatology organisations and charity funders following suit (8). However, whereas integration of PPI in clinical research in rheumatology is now widespread, implementing meaningful PPI in pre-clinical, discovery science research has proven more challenging (9). Discovery scientists’ concerns around integrating PPI in their research are varied and include: problems communicating complex scientific information to the lay public, concerns around public opinion on animal testing, lack of knowledge around how they can implement PPI in their research, and the time and resources needed to deliver successful and impactful PPI (2). Furthermore, patients have indicated concerns about difficulty in understanding the message conveyed by discovery scientists and that they feel they do not have a common language (10), although there have been examples of effective solutions, such as the co-development of a glossary of relevant scientific terms and concepts (11).

PPI in discovery science is an important means to enhance research accountability, ensuring that research is relevant to the needs and priorities of the target community, and to increase researcher motivation by better understanding the impact of the conditions they are researching. PPI contributors can further review and improve lay materials in grant applications and, where applicable, patient-facing materials, and be involved in the dissemination activities and advocacy roles (8, 12).

In order to facilitate the integration of PPI in discovery research, relevant training for both researchers and PPI contributors should be considered (9). A report of a meeting of PPI contributors and researchers involved in the Research into Inflammatory Arthritis Centre Versus Arthritis (RACE) highlighted patient partners’ views that PPI should be integrated into researchers’ training from the earliest stages to facilitate meaningful patient involvement in rheumatology research (10).

A few examples of successful integration of PPI in doctoral training in other disease areas have described positive impacts of the collaboration with PPI partners such as improved researcher self-esteem, reduced student isolation, a beneficial impact on the progression of research and an increased understanding of how PPI can be integrated into research (e.g., (13-15), but further examples are needed, especially in relation to discovery science.

Recognising the need for PhD researchers to understand the value of PPI, to learn about patient experiences of disease and priorities for research, and to learn how to communicate with the public about their research, the Student Patient Alliance (SPA) initiative was developed by the Rheumatology Research Group (RRG) and the Rheumatology Research Patient Partnership (R2P2 (16)) at the University of Birmingham, UK. The SPA initiative partners doctoral research students, the majority of whom are engaged in pre-clinical research, with one or more PPI contributors throughout their doctoral studies and provides training and information resources for both students and their PPI partners for this process.

The SPA has now been implemented across collaborating research centres and this paper describes the development and evaluation of the SPA from the perspectives of the students, their PPI partners and PPI coordinators in the participating centres. We aim to share our experiences and reflect on the findings of the evaluation to identify barriers and facilitators to effective partnerships between doctoral students and PPI contributors, which could be considered in future initiatives of this kind.

## METHODS

The SPA was developed in two phases, starting with a local pilot which ran from December 2018 to June 2020, and a full implementation phase across multiple research centres and collaborating sites in early 2021. The feedback and experiences from the pilot were used as a basis for the implementation of the SPA on a larger scale and informed the materials supplied.

### Student Patient Alliance initial pilot scheme

The initial pilot of the SPA started in December 2018 at the University of Birmingham with three PhD students undertaking laboratory-based rheumatology research, and seven PPI contributors from R2P2 (all of whom were patients with rheumatic conditions). Students were partnered with one or more PPI contributors who had volunteered for the SPA by the R2P2 coordinators based on the students’ research area and the interests of the PPI contributors and, where possible, the relevant disease area. During an initial face-to-face ‘kick-off’ meeting, both groups received further information about the SPA and were provided with an opportunity to introduce themselves to each other informally.

Students and their PPI partners were given information about PPI and related training resources and opportunities, including guidance on writing/providing feedback on a lay summary of research. Students were subsequently asked to take the lead in contacting their PPI partners and facilitating their involvement. They were asked to conduct at least one PPI activity with their partner(s) (e.g., producing a poster or an oral presentation for the general public). A list of the actions required of the students is shown in Table 1.

**Table 1.**
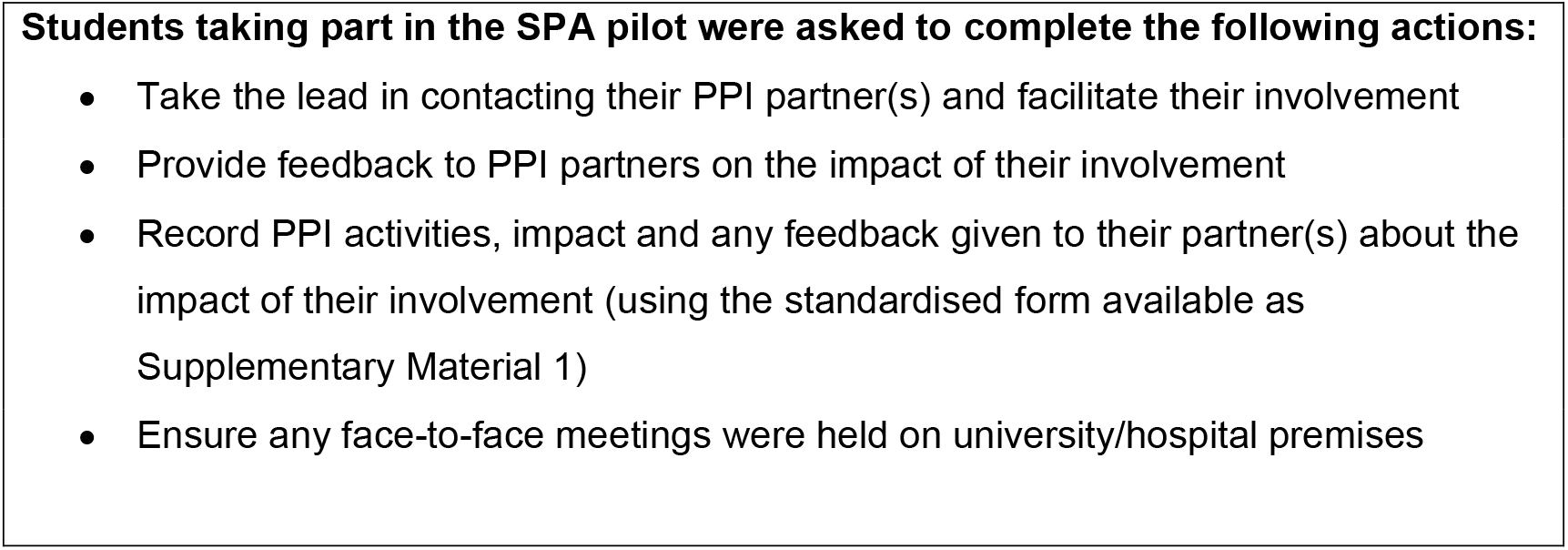
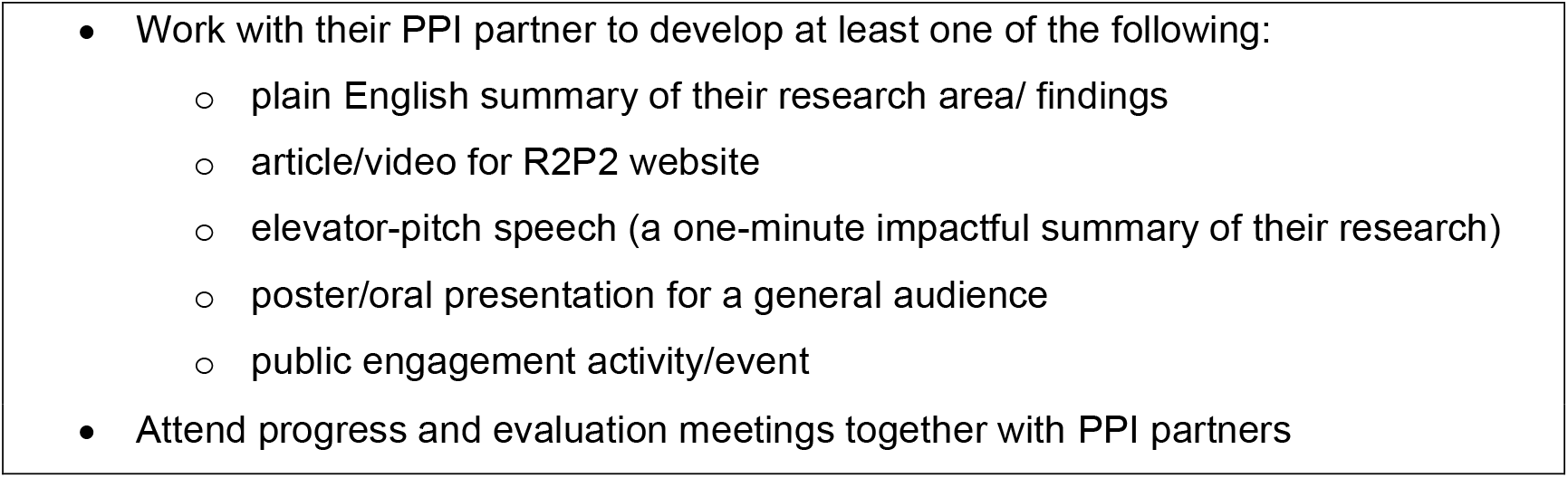
SPA local pilot – expectation for student participation

As part of the pilot, all three students attended a progress meeting with GS and RB in April 2019. The students and four of the PPI partners attended a follow-up meeting with MF, GS and RB in February 2020 to discuss their experiences of working together and identify issues to be addressed in further iterations of the SPA. This discussion was audio recorded and transcribed verbatim by RB. The meeting transcript was read by MF and GS who independently noted salient discussion points and related quotations, and then met to compare and collate them into a narrative description.

### Student Patient Alliance implementation across collaborating centres

After a delay of approximately 2 months, as a result of the developing COVID-19 pandemic, the SPA was implemented (online) in January-February 2021 for rheumatology research doctoral students at the University of Birmingham and across partnering institutions in our centres of excellence (RACE, funded by Versus Arthritis, incorporating the Universities of Birmingham, Glasgow, Newcastle and Oxford and the Centre for Musculoskeletal Ageing Research (CMAR), funded by Versus Arthritis and the Medical Research Council which includes the Universities of Birmingham and Nottingham).

The PPI coordinators at each research centre invited PPI contributors from the local PPI groups to participate in the SPA initiative, matched students with the volunteer PPI contributors and facilitated initial online meetings. The matching of students and PPI contributors varied across centres and sites. Students typically produced a short lay summary of their intended or ongoing PhD research project which was either distributed to the PPI contributors before an initial online meeting or presented by the students to the PPI contributors during this introductory meeting. Where possible, PPI coordinators matched students and PPI contributors with similar interests and within the relevant disease area. In several cases, the PPI contributors were given the opportunity (either in a type of ‘speed dating’ exercise during the meeting or via email prior to the meeting) to indicate the PhD project they were most interested in, and where possible a match was made on that basis.

Information packs and resources for students and their PPI partners used in the pilot scheme were updated to reflect learnings from the SPA pilot exercise and to include additional guidance for remote meetings between students and their PPI partners in response to the COVID-19 pandemic. These materials were provided to the designated PPI coordinators at each participating centre, who were asked to adapt these for their local context and distribute them to students and their PPI partners, following the procedures used for the pilot scheme at the University of Birmingham. PPI coordinators were also asked to remind students to complete a record of PPI activity (Supplementary Materials 2). Students were further given the same list of expectations as in the pilot (Table 1) with the additional instruction to move meetings to Zoom/Teams and/or communicate via phone/ email when requested by the PPI partner or when necessary due to the developing pandemic.

Two feedback questionnaires (one for students and another for PPI contributors) were developed by the PPI team at Birmingham, with input from students and PPI partners participating in the SPA pilot scheme who completed and returned a draft version of the questionnaires and provided feedback on its content and format. This feedback was incorporated in the final web-based version of the questionnaires (Supplementary Materials 3). All participating students and PPI partners were asked by their local PPI coordinators to complete the relevant questionnaire approximately six months after starting with the SPA. Both questionnaires included several statements regarding impact to which respondents indicated their level of agreement on a 5-point Likert scale, as well as several open-ended feedback questions. PPI partners were further asked which research activities they were involved in with their current SPA student. The PPI partner feedback questionnaire also included a 10-point Likert scale measuring their satisfaction with the collaboration with their student partner(s) (1 is not at all satisfied, 10 is extremely satisfied).

In addition, in November 2021, the PPI coordinators at each participating centre were asked to complete a short survey on their experiences of the SPA scheme. In a series of open-ended questions, they were asked if they shared the information pack with students and/or PPI contributors and if they modified any of the documents and/or provided additional resources, as well as questions about the local matching process and support they had given to their students and/or PPI partners.

## RESULTS

### Learnings from the SPA pilot

During the pilot, all three students collaborated with their PPI partner(s) to co-produce outputs and activities including award-winning posters, published online abstracts (17), exhibits for public engagement events at the University of Birmingham and oral co-presentations at an annual meeting of the R2P2 patient partnership group. Specific opportunities for collaborations of this kind were reported as useful to structure meetings between students and patients. Quotes from the pilot progress meeting are inserted below and can be found in Supplementary Materials 4.

All students and their PPI partners described their experience of working together as very positive. In addition to the development of public engagement skills, students reported that explaining their research to their partners at an early stage in their project helped to consolidate their own understanding of their field. Students further valued learning from patients’ experiences of illness and treatment, which increased their understanding of the clinical relevance of their research and enhanced their own motivation:

*“It kept my focus on the bench-to-bedside aspect because I think us being basic science researchers, we’re really focused on getting the right research design, having the right experiments, testing everything at a very molecular level and I think we tend to forget that the only reason that we’re doing this is to actually improve patients’ lives… And it does bring my motivation up quite a bit, particularly if things aren’t going terribly well in the labs or if I’m in a transition part of the project*.*”* (Student)

PPI partners reported enjoying learning about innovative scientific research and hearing about research progress, which they described as inspiring and providing hope for the future. They also enjoyed providing positive feedback and encouragement to the students, which students in turn described as motivating.

*“I could see, yes you were getting somewhere with it and there’s almost a feeling of excitement that when we next meet up in the next couple of months or so - I’m interested to see, has that line of research gone further. And maybe with everybody that’s studying and working its quite good to have perhaps some positive feedback and I think the patient partners can do that. Because you do feel this gratitude for the fact that somebody’s doing something that might help you but probably more likely help other people*.*”* (PPI partner)

When discussing areas to address in future initiatives, it was noted that not all students would be proactive or comfortable about leading the collaboration and it was suggested to highlight key benefits, for both students and patients, at the outset.

However, it was felt that the student-led approach offered flexibility and informality that fostered relationship-building between students and patients. Students and their PPI partners agreed that some potential PPI partners may not be comfortable with supporting research involving animals and highlighted the importance of making PPI partners aware of any potential animal work before students and their partners begin working together. The findings from the pilot were used to inform the methodology and formal implementation of SPA across collaborative research centres. The SPA feedback questionnaires were updated with input from pilot students and PPI partners and additional materials were made available for the new cohorts, including a video about working together to produce research posters made by a student and their PPI partner from the SPA pilot. Feedback around issues such as animal work further informed introductory meetings.

### Full SPA implementation across collaborating centres

#### Student’s and PPI partner’s characteristics

For the first full implementation of the SPA, a total of 20 PhD students took part from across the CMAR and RACE research centres, of whom 19 students completed the feedback forms. One student had taken a leave of absence at the time of survey completion. Seven of the 19 students (37%) were in their first year of study, six (32%) in their second year and the remaining six (32%) in their third year of study. Fifteen students (79%) were entirely laboratory-based, two were doing clinical research and two combined laboratory and clinical research.

A total of 20 PPI partners were originally involved with this implementation of the SPA, although two contributors had to drop out for unspecified reasons (see also Table 2). The majority of students were teamed with one PPI partner each, one student had two PPI partners and one student had no PPI support due to the drop-out of their PPI partner (a new PPI partner has since been found, but data are not yet available). One PPI partner contributor supported two students at different CMAR sites. Of the 18 PPI partners who stayed in the programme, 16 completed the feedback forms, with one completing feedback forms for both of their students. Eight students (42%) indicated that they worked with a PPI partner(s) who had a disease they perceived to be directly relevant to their research area. Further details of the student participants and their partners across the centres can be found in Table 2.

**Table 2.** Student and PPI partners per centre and site

|  | **Students**<br>**total** | **Lab-**<br>**based**<br>**PhD** | **Clinical**<br>**PhD** | **Clinical**<br>**and lab-**<br>**based**<br>**PhD** | **Number of**<br>**PPI**<br>**contributors**<br>**assigned**<br>**per centre** | **Number of PPI**<br>**contributors with**<br>**relevant disease** |
| --- | --- | --- | --- | --- | --- | --- |
| **Centre and site** | **N** | **N** | **N** | **N** | **N** | **N** |
| CMAR Birmingham | 7 | 7 |  |  | 7* | 1 |
| CMAR Nottingham | 6 | 2 | 2 | 2 | 6* | 2 |
| RACE Birmingham | 1 | 1 |  |  | 1 | 1 |
| RACE Glasgow | 2 | 2 |  |  | 4** | 2 |
| RACE Newcastle | 1 | 1 |  |  | 1 | 1 |
| RACE Oxford | 2 | 2 |  |  | 2** | 2 |
| *Total* | *19* | *15* | *2* | *2* | *20* | *9* |

\* One PPI contributor supported a student at each of the CMAR sites; \*\* One PPI contributor dropped out at these sites.

Nine students (47%) first learned about the SPA initiative from the local PPI coordinator or other university staff; eight (42%) from their PhD supervisor and the remainder from another source. Eleven PPI partners (65%) first learned about the SPA initiative from their local PPI group, the others heard about it through a community notice board, email or website, word of mouth or from University or PPI staff.

#### Student feedback related to SPA & PPI resources, training and support

Seventeen of the 19 students who completed the survey (89%) reported they had received PPI resources. Fourteen reported that they had received the dedicated SPA resources and five of these students also received NIHR/INVOLVE guidance and/ or research funder resources. Three students stated they had only received PPI information from their research funder. Fifteen (88%) of the 17 students who received PPI resources found them useful (Table 3 gives an overview of the students’ feedback evaluation).

**Table 3.** Student SPA evaluation scoring (closed questions)

| Statements | Rating |  |  |  |  |  |
| --- | --- | --- | --- | --- | --- | --- |
|  | Not at all useful | Not useful | Neutral | Useful | Very Useful | n/a |
| SPA & PPI resources / training / support (N = 19) | N (%) | N (%) | N (%) | N (%) | N (%) | N (%) |
| How useful were the resources you received? | 0 | 1 (5) | 2 (10) | 9 (47) | 4 (21) | 3 (16) |
| Impact (N = 19) | Strongly Disagree | Disagree | Neutral | Agree | Strongly Agree | n/a |
|  | N (%) | N (%) | N (%) | N (%) | N (%) | N (%) |
| SPA improved my knowledge about patient/public involvement in research. | 0 | 1 (5) | 4 (21) | 10 (53) | 4 (21) | 0 |
| SPA improved my confidence to communicate about my research with members of the public. | 0 | 1 (5) | 2 (10) | 11 (58) | 5 (26) | 0 |
| SPA changed aspects of my research design. | 5 (26) | 7 (37) | 5 (26) | 1 (5) | 0 | 1 (5) |
| SPA increased my interest in my research area. | 1 (5) | 2 (10) | 9 (47) | 6 (32) | 1 (5) | 0 |
| SPA shaped my future research/career plans. | 3 (16) | 5 (26) | 10 (53) | 1 (5) | 0 | 0 |
| SPA improved my understanding of research ethics. | 1(5) | 4 (21) | 7 (37) | 7 (37) | 0 | 0 |
| SPA improved my knowledge of patient/public priorities for research | 0 | 1(5) | 4 (21) | 9 (47) | 5 (26) | 0 |
| **Overall impact (N = 19)** | **Very negative impact** | **Negative impact** | **Neither negative nor positive** | **Positive impact** | **Very positive impact** | **n/a** |
|  | **N (%)** | **N (%)** | **N (%)** | **N (%)** | **N (%)** | **N (%)** |
| Extent to which collaboration with patient/public research partner(s) had a positive or negative impact on you and your research. | 0 | 0 | 8 (42) | 9 (47) | 2 (10) | 0 |
| **Understanding (N = 18)** | **Strongly Disagree** | **Disagree** | **Neutral** | **Agree** | **Strongly Agree** | **n/a** |
|  | **N (%)** | **N (%)** | **N (%)** | **N (%)** | **N (%)** | **N (%)** |
| Taking part in the SPA has improved my understanding of my partner's disease.* | 0 | 0 | 1 (6) | 5 (28) | 1 (6) | 11 (61)** |
| Taking part in the SPA has improved my understanding of my research from the public's point of view.* | 0 | 0 | 3 (17) | 11 (61) | 4 (22) | 0 |

| Recommendation (N = 19) | Not very likely | Not likely | Neither likely nor unlikely | Likely | Very likely | n/a |
| --- | --- | --- | --- | --- | --- | --- |
|  | N (%) | N (%) | N (%) | N (%) | N (%) | N (%) |
| How likely are you to recommend the SPA to other research students? | 0 | 2 (10) | 6(32) | 7(37) | 4(21) | 0 |

Please note, the responses for one student were based on their experience of collaborating with two PPI contributors, but they completed one survey. \*One student could not complete the 'Understanding' questions as they had had no contact with their allocated PPI partner, so frequencies are based on 18 responses for those questions. \*\*This includes students who stated that their PPI partner did not have a disease relevant to their research area.

The students also made suggestions for resources and training they felt would have helped them, including free access to the Pro version of Zoom for all students and their PPI partners to facilitate online meetings without time limits during the pandemic when in-person meetings were not possible (this was available to students at some universities but not others). Other resources they would have found useful included examples of a good poster or presentation in plain English, and information on conveying scientific methods to a public audience, demonstrating appropriate content and language to use. It was also suggested by some that students and PPI partners should be linked up earlier in the PhD cycle (due to the timing of the SPA roll-out, this was only later in their PhD for some students).

In terms of additional support, 12 of the students (63%) indicated that they received some form of administrative support for PPI from their host institution in the form of arranging telephone or video calls with PPI partners; providing information about PPI and training opportunities; support with reimbursement of PPI partners’ expenses and payments.

#### Student feedback related to the impact of the SPA

Fourteen students (74%) agreed or strongly agreed that participating in the SPA improved their knowledge of PPI and patient priorities and 16 (84%) indicated that it increased their confidence in communicating with the public, but many did not agree with the statement that the SPA changed their research design (*n* =12; 63% disagreed/strongly disagreed) or their future research plans (*n* = 8; 42% disagreed/strongly disagreed) as a result of their interactions with their PPI partner. Students’ evaluation of the impact of SPA is summarised in Table 3.

Fifteen students (79%) further described positive impacts of taking part in SPA in the free text responses. They described how engaging with their PPI partner improved their communication and public engagement skills; how they found it beneficial to hear from people with a chronic condition and how it affects daily life; and that they gained new perspectives.

*“I feel more confident speaking about immunology, especially to someone with less/no experience in the field. I have thoroughly enjoyed trying to answer my patient’s questions and has helped me understand the immunology from a clinical perspective with ‘real-world’ examples*.*”*

Five students (26%) also reported negative impacts or challenges in their free-text responses. These included: difficulty in making laboratory research interesting for PPI partners; challenges in finding appropriate activities for PPI partners; and reduced or delayed opportunities to engage with PPI partners due to the COVID-19 pandemic. Whilst some students identified time commitment as a negatively impacting factor, others recognised the time commitment as a potential negative impact but described it as minimal, or felt it was worth the time and effort:

*“I wouldn’t say being part of the SPA programme has had any significant negative impact on me or my research overall. It takes a bit of time, but I think what I get out of it is absolutely worth the extra time I put into it*.*”*

Furthermore, 6 out of 8 students (75%) who reported that their partner(s) had a health condition directly relevant to their research area, agreed or strongly agreed that taking part in the SPA had improved their understanding of this disease and 15 students (83%) (strongly) agreed that taking part in the SPA had improved their understanding of their research from the public’s point of view. Eleven (61%) would be likely or very likely to recommend the SPA to others (see also Table 3).

There were no apparent differences in feedback between those students who were partnered with a PPI partner whom they perceived to have a relevant illness to their PhD and those that perceived this was not the case in any of the ratings. There were also no apparent differences between those students who were purely laboratory-based and those who had a clinical element to their research.

#### PPI partners’ feedback related to involvement in research activities

Table 4 gives an overview of PPI partner involvement in specific research activities in collaboration with their SPA student. Seven PPI partners (41%) reported being involved in developing plain English summaries of research and nine (53%) were involved in the development of research presentations and posters. Four PPI partners (23%) reported being involved in advising on research design, analysis or findings. Nine PPI partners (53%) were involved in at least two research activities during their collaboration with the student(s).

**Table 4.** PPI partners participation in specific research activities as part of their collaboration with their student partner.

| **Research activity (N = 17)** | **N (%)** |
| --- | --- |
| Advising on research design/analysis/findings | 4 (23) |
| Advising on research documents (e.g., consent forms/ information for participants) | 2 (12) |
| Contributing to public events about research; | 1 (6) |
| Developing plain English summaries of research | 7 (41) |
| Developing research presentations/posters | 9 (53) |
| Developing research priorities/ideas | 1 (6) |
| Other research activities (e.g., elevator speech; advised on participant recruitment, etc) | 6 (35) |

Please note, one PPI contributor completed 2 evaluation surveys, 1 for each of the students they were partnered with and the data from both evaluation surveys is included in this table.

#### PPI partners’ feedback on interactions with their student partner(s) and the impact of the collaboration

Most PPI partners were (extremely) satisfied with the amount of interaction they had with their student partner(s) and a majority of the PPI contributors felt they received enough feedback from their student partner(s) on the impact of their involvement in the research project(s). Table 5 gives an overview of the PPI partners’ feedback scoring. Most PPI partners (N = 14; 82%) indicated that overall, the collaboration with their student partner(s) had a positive/very positive impact on them, with none reporting a negative impact (see Table 5). PPI contributors further described specific positive aspects such as having “friendly, fruitful and meaningful” discussions with their student, being able to pass on the benefit of their experiences and being able to put their coaching and mentoring skills into practice, amongst other things. They looked forward to continuing the partnership:

**Table 5.**
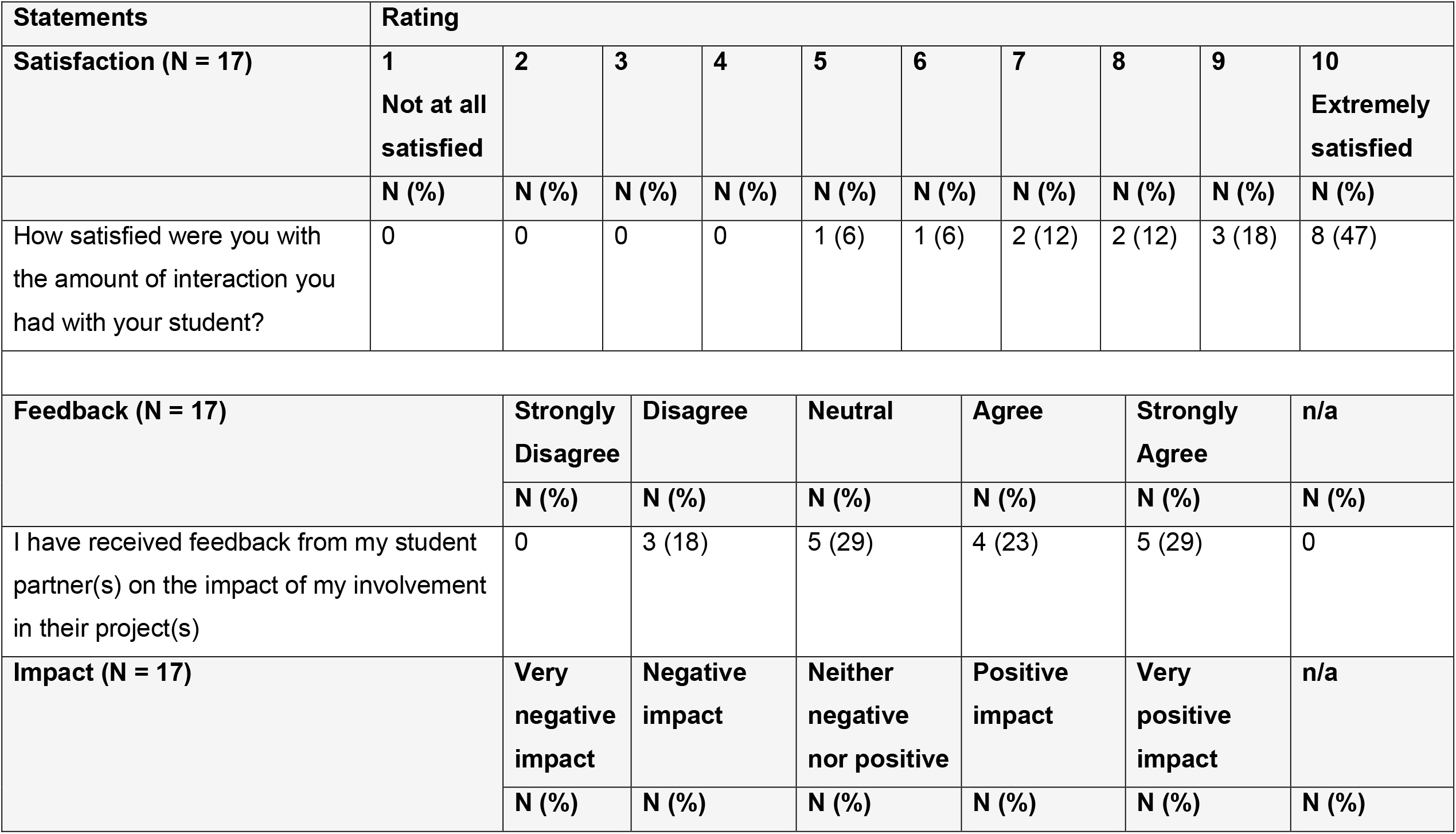
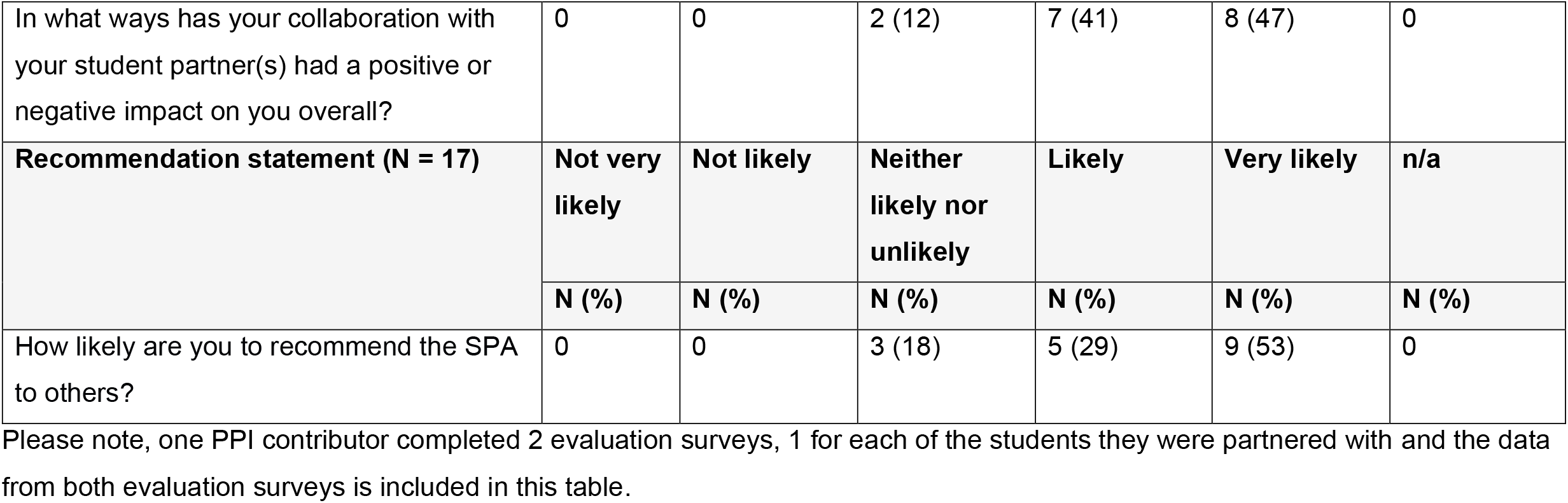
PPI partner SPA evaluation (closed questions)

| Statements | Rating |  |  |  |  |  |  |  |  |  |
| --- | --- | --- | --- | --- | --- | --- | --- | --- | --- | --- |
| Satisfaction (N = 17) | 1<br>Not at all<br>satisfied | 2 | 3 | 4 | 5 | 6 | 7 | 8 | 9 | 10<br>Extremely<br>satisfied |
|  | N (%) | N (%) | N (%) | N (%) | N (%) | N (%) | N (%) | N (%) | N (%) | N (%) |
| How satisfied were you with the amount of interaction you had with your student? | 0 | 0 | 0 | 0 | 1 (6) | 1 (6) | 2 (12) | 2 (12) | 3 (18) | 8 (47) |
| Feedback (N = 17) | Strongly<br>Disagree | Disagree | Neutral | Agree | Strongly<br>Agree | n/a |  |  |  |  |
|  | N (%) | N (%) | N (%) | N (%) | N (%) | N (%) |  |  |  |  |
| I have received feedback from my student partner(s) on the impact of my involvement in their project(s) | 0 | 3 (18) | 5 (29) | 4 (23) | 5 (29) | 0 |  |  |  |  |
| Impact (N = 17) | Very<br>negative<br>impact | Negative<br>impact | Neither<br>negative<br>nor positive | Positive<br>impact | Very<br>positive<br>impact | n/a |  |  |  |  |
|  | N (%) | N (%) | N (%) | N (%) | N (%) | N (%) |  |  |  |  |

|  |  |  |  |  |  |  |
| --- | --- | --- | --- | --- | --- | --- |
| In what ways has your collaboration with your student partner(s) had a positive or negative impact on you overall? | 0 | 0 | 2 (12) | 7 (41) | 8 (47) | 0 |
| **Recommendation statement (N = 17)** | **Not very likely** | **Not likely** | **Neither likely nor unlikely** | **Likely** | **Very likely** | **n/a** |
|  | **N (%)** | **N (%)** | **N (%)** | **N (%)** | **N (%)** | **N (%)** |
| How likely are you to recommend the SPA to others? | 0 | 0 | 3 (18) | 5 (29) | 9 (53) | 0 |

Please note, one PPI contributor completed 2 evaluation surveys, 1 for each of the students they were partnered with and the data from both evaluation surveys is included in this table.

*“This is currently an ongoing process and I look forward to supporting my student for the duration of* [their] *research. I felt the initial sessions over Zoom were well organised to introduce the opportunities to volunteers like myself and introductions to the students enabled a clear understanding of this role to be realised. Matching students and volunteers was well thought out*.*”* (PPI partner)

PPI contributors also perceived some negative aspects. For example, one individual felt that within the overall SPA programme, there was a lack of equality, diversity and inclusion in terms of university staff and student representation. Another PPI contributor mentioned that they did not feel that their student partner’s supervisor understood the role of PPI in research. Some perceived the student projects to be pre-defined and felt this made it less of a PPI opportunity:

*“In my opinion, this is not true PPI project as the research is already decided at this stage of the academic life cycle, with very specific outcomes*.” (PPI partner)

The fact that no in-person meetings could take place due to COVID-19 was also perceived negatively. However, the majority of PPI partners (N = 13; 76%) were likely or very likely to recommend the SPA scheme to others, with none not likely to recommend the SPA (see Table 5).

#### PPI partners’ feedback on training and learning opportunities

All but one of the PPI partners who completed the survey indicated that they had undertaken/received a training or learning opportunity during their involvement with the SPA. In most cases this involved courses or workshops about PPI in research, often in combination with the use of the SPA and/or INVOLVE/NIHR materials provided. PPI partners indicated that in future they would like to receive information on who to contact when a student was not in regular contact. They would also like support with improving virtual interactions and would like more in-person seminars and workshops.

#### Feedback from the PPI coordinators at the research centres

Feedback was received from three PPI coordinators who implemented the SPA in their local teams. They reported they shared the documents in the information pack provided by Birmingham with the students and PPI contributors although some modifications were made to ensure the documents were relevant to the local research centre/ site. Not all available information related to virtual meetings during the pandemic was shared as different sites used different platforms.

Coordinators indicated that it was not always possible to match the student with a PPI partner with a health condition directly relevant to their PhD research topic and that students had been informed in advance that this was a possibility. Where appropriate, PPI partners were also advised that there was a possibility that the PhD topic might not match their particular health condition or that they might not be matched with the student whose project they found most interesting.

PPI coordinators were positive about SPA. For example, one indicated that they felt that the SPA initiative was important in developing the researchers of the future and that PPI should be embedded from the very start of the students’ research careers. Others indicated that they felt students benefited from the SPA in terms of seeing their research from a new perspective and being able to communicate their ideas to new audiences. Furthermore, it was felt that the PPI partners benefitted socially from being involved in the SPA and valued feeling they were giving something back in return for the medical care they had received in the past.

PPI coordinators had several recommendations for future iterations of the SPA. They indicated that at a local level, it would be useful to supplement generic SPA resources with other in-house opportunities, such as local PPI workshops. They also felt it was important to have a contingency plan in place in case students or PPI partners were unable to continue with the SPA and to be clearer about this possibility with all parties from the outset. They also indicated that the recruitment of PPI contributors for the SPA should start early for new cohorts of students, but they would use similar processes to match/introduce students to PPI partners. They further highlighted the importance of managing expectations of students and their PPI partners around the matching process but also the type of activities the students and their partners would be likely to undertake together from the outset.

## DISCUSSION

Our study provides one of few accounts of the development and evaluation of an initiative to partner doctoral students (predominantly engaged in discovery, laboratory-based science) with PPI partners. Overall, the pilot and first wave of full implementation of the SPA have been a successful and largely positive experience as evidenced by the feedback from participants, both students and PPI partners. In addition, some important lessons have been learned through the implementation and evaluation, which should lead to further improvements for future iterations of this SPA.

Both students and their PPI partners felt that taking part in SPA had a positive or very positive impact. Students reported an increased understanding of PPI and patient priorities and reported improved public engagement and communication skills. They valued getting different perspectives and learning how chronic health conditions may affect the daily lives of those who live with these conditions. PPI partners highlighted the positive impact of having meaningful discussions and acting as a mentor to ‘their’ students.

Most students indicated that they did not change their research design or their current and future research plans as a result of PPI, though this was not an explicit objective of the SPA initiative. This also reflects that many of the students had already started their PhD when the SPA was rolled out and as a consequence the opportunities to change the design or future plans were more limited. In some centres however, PPI partners were involved from an earlier stage in the selection of PhD student projects. Challenges reported by students included the time commitment needed for effective PPI and difficulty finding appropriate involvement opportunities for their PPI partners. Both students and PPI contributors further mentioned limitations due to the COVID-19 pandemic and expressed the hope for future face-to-face meetings, recognising an important social component of the SPA partnership. Indeed, feelings of loneliness and anxiety as a result of the enforced social isolation during COVID related lockdowns have been found to affect large proportions of academic staff and students (e.g.(18))

Findings from previous research and evaluations around the integration of PPI in doctoral studies in other areas have also been largely positive (14, 15). The current findings are in line with findings that PPI contributed to motivating junior discovery scientists and was valued by both researchers and PPI partners, though PPI was not perceived to impact the discovery science directly (9). In part, this might be the nature of PhD research projects in the biomedical sciences, where the doctoral candidate’s main research theme and programme are often predefined as part of the initial funding application. One PPI partner in the present study felt that the PhD supervisor did not value PPI and thus this collaboration might have suffered as a result. This highlights the need for embedding PPI at an early career stage using methods as exemplified by the SPA, and the importance of support from senior investigators.

Although it did not appear to impact the student ratings in the relatively small sample of this evaluation, not all students felt that they were matched up with a PPI partner who had a disease relevant to their PhD topic. Some PPI coordinators indicated that they struggled to find PPI partners with the relevant condition for some students and suggested it would be prudent to start the search for PPI partners at an earlier stage, although the positive experiences in our study suggest this need not be an absolute requirement.

Students indicated a desire for further training materials addressing approaches to conveying scientific methods to a lay audience, including the appropriate content and language to use, as well as examples of these. Although training resources were made available and included a guide to writing a lay summary for both researchers and PPI partners, it appears these were not always received, not perceived to be comprehensive enough or not relevant to the needs of students doing complex discovery science research.

Although providing feedback to patient partners was a requirement both in the SPA pilot and full implementation phase, this did not happen in all collaborations. Feedback enables PPI partners to assess whether their input has been valued, and impactful. It also motivates them and increases their confidence (19). To retain PPI contributors for the SPA and similar initiatives, more formal requirements/expectations may be needed to ensure effective feedback to PPI partners. Here there may be a role for PPI coordinators, as well as support from students’ supervisors. We did not formally seek feedback from the students’ supervisors in this study, but this would be helpful in future to understand their perceptions of the SPA on the student and their research, and also to assess whether this changes the supervisors’ views and interactions with PPI.

We hope that the findings of this evaluation of the SPA could inform the implementation of similar PPI programmes for PhD students. Specifically, these should include the provision of a core set of information resources describing what PPI is and how the SPA (or similar scheme) works. It should further include information about conducting virtual meetings via a variety of platforms and guidance for in-person meetings (including any relevant safeguarding measures), practical support for payment for PPI contributors and some training materials on writing lay summaries and communicating to a lay audience. Offering further training through workshops to students, PhD supervisors and PPI contributors is also likely to be beneficial. In addition to formal resources, our results indicate that clear descriptions of expectations from the outset are important for all parties. Dedicated support staff with PPI expertise and experience are likely to be beneficial to any scheme that integrates PPI into doctoral training programmes. Finally, resources and infrastructure to support the availability of a large and diverse pool of PPI contributors, ideally with a relevant condition or with lived experience (i.e., as a patient or carer) of the relevant conditions could facilitate matching the research area of interest and the PPI contributor’s health condition and interests, where relevant. Where patient partner availability is limited, partnering a patient with a group of students may be effective. Although it is likely that individual partnerships facilitate relationship building and confidence for junior researchers, to the authors’ knowledge, no studies have directly compared group and individual partnership approaches.

Limitations of the current evaluation of the SPA include the small sample size. As such, it was not possible to systematically assess variations across centres. For example, it was more difficult to link students up with a PPI partner with a disease they considered relevant at some sites than it was at those centres supported by established disease-specific PPI groups. Some students’ projects were focused on musculoskeletal ageing/sarcopenia and not on specific diseases such as rheumatoid arthritis (RA). Whilst some centres had access to PPI groups which included significant numbers of patients with specific conditions, others were supported by PPI groups that included healthy older members of the public. However, the current findings suggest that this did not have a negative impact on the extent to which partnerships between students and PPI partners were valued.

## Conclusions

In conclusion, this account of our experiences with the SPA demonstrates a wealth of positive impacts for students and interested PPI partners, and highlights the resources needed to facilitate effective partnerships and implementation on a wider scale, such as dedicated staff with PPI expertise, tailored training opportunities, infrastructure to support access to a large group of patient partners, and active support from PhD supervisors.

## Data Availability

The datasets generated and analysed during the current study are not publicly available due to the small sample size and the likelihood that individual data could be associated with a specific person. SPA resources are available upon application to the senior author.

## List of abbreviations

CMAR: MRC-Versus Arthritis Centre for Musculoskeletal Ageing Research
COVID-19: Coronavirus Disease 2019
EULAR: the European Alliance of Associations for Rheumatology
NIHR: National Institute for Health Research
PhD: Doctor of Philosophy
PPI: Patient and Public Involvement
RACE: Research into inflammatory Arthritis Centre Versus Arthritis
R2P2: Rheumatology Research Patient Partnership
SPA: Student Patient Alliance

## DECLARATIONS

### Ethics approval and consent to participate

In this paper, we report on the development and implementation of the Student Patient Alliance. Patient Research Partners and students were involved in the design of SPA and its evaluation. All participants were provided with information about the purpose of the evaluation. Participants verbally agreed to the recording of the evaluation meeting of the pilot programme. Completion and return of any of the surveys was voluntary for all and implied consent to participate. This was a service evaluation and according to the UK Health Research Authority, formal ethical approval is not needed for research of this kind.

### Consent for publication

Not applicable; any quotes taken from the surveys are not associated with an individual.

### Authors’ contributions

GS, KR, MF and RB have made substantial contributions to the conception and design of the work. EI, RR, GN have been involved in the pilot of the work and further design of the work. GS, RB, BEP, HMC, LR, CW, SWJ, AGP SS, KR and MF have been involved in the full implementation of the work; GS, RB, HMC, LR, CW, SWJ, KR and MF have contributed to the acquisition, analysis interpretation of data; GS, RB, BEP, AGP SS JS KR and MF have drafted the work or substantively revised it. All authors have read and approved the submitted version of the manuscript.

## Acknowledgements

This work was supported by the Research into Inflammatory Arthritis Centre Versus Arthritis (RACE) (grant number 22072).

We are immensely grateful to all PPI contributors, doctoral students and PPI coordinators who have taken part in the SPA pilot and the first full-scale implementation of the SPA. In particular we would like to thank Laura Chapman, Jonathan Lewis, Julia Manning, Linda Murphy, Caroline Morris, Janet Munro, and Jackie Erpen. We are also grateful for support from the Research into Inflammatory Arthritis Centre Versus Arthritis, at the Universities of Birmingham, Glasgow, Newcastle and Oxford.

