## Supplemental Materials for "The Student Patient Alliance: Development and formative evaluation of an initiative to support collaborations between patient and public involvement contributors and doctoral students"

### Supplementary materials

1. Feedback forms students and PPI partners used in the pilot: pages 2-5
2. Student log to record PPI activities: page 6
3. Web-based feedback forms for students and PPI partners: pages 7-13
4. Quotes pilot SPA evaluation: pages 14-15

Supplementary Materials 1: Feedback forms pilot

**Rheumatology Research Patient Partnership (R2P2)**

**Student Patient Alliance (SPA) – Student Feedback Form**

| | | |
|---|---|---|
| **Student name:** | | **Patient partner name(s):** |

Please complete the following table to record in detail your patient/public involvement activities. Insert additional rows whenever necessary.

| **Purpose of activity** (E.g. develop lay summary/news article, discuss research priorities…) | **Type of activity** (E.g. face to face meeting; email exchange…) | **Who initiated the activity?** (E.g. student, patient partner, supervisor, PPI lead…) | **Date(s) of Activity** | **Impact of activity** (E.g. changes made to research design/documents; things you have learnt, skills you have developed as a result of the interaction) | **Date(s) feedback on impact of activity given to patient partner** | **Type of feedback** (E.g. email message, phone call) | **Comments** (E.g. challenges encountered, solutions implemented, training needs identified, future plans…) |
|---|---|---|---|---|---|---|---|

| |
|---|
| When you met your patient partner(s), you were given some guidance and information about patient involvement in research. Please describe any additional resources/information that would have been useful: |
| In what ways has your collaboration with your patient research partner(s) had a positive impact on you / your research? |
| In what ways has your collaboration with your patient research partner(s) had a negative impact on you / your research? |
| Please suggest aspects of the Student Patient Alliance that could be improved: |
| Please describe any additional thoughts about the Student Patient Alliance that have not been captured in this form: |

**Thank you!**

**Rheumatology Research Patient Partnership (R2P2)**

**Student Patient Alliance (SPA) – Patient Partner Feedback Form**

| Your name: |
|---|
| Date: |
| Student partner's name: |
| Please describe any interactions you have had with your student partner: |
| Please circle a number to indicate how satisfied you are with the amount of interaction that you have had with your student partner?<br><br>0 1 2 3 4 5 6 7 8 9 10<br>Not at all satisfied … Extremely satisfied<br><br>Please explain your answer: |
| Please circle a number to indicate how satisfied you are with the feedback that you have received from your student partner about the impact of your involvement on their research?<br><br>0 1 2 3 4 5 6 7 8 9 10<br>Not at all satisfied … Extremely satisfied<br><br>Please explain your answer: |
| In what ways has your collaboration with your student research partner had a positive impact on you? |
| In what ways has your collaboration with your student research partner had a negative impact on you? |
| In what ways has your collaboration with your student research partner had a positive impact on the student and their project? |
| In what ways has your collaboration with your student research partner had a negative impact on the student and their project? |

| |
|---|
| Have any expenses you have incurred as a result of your collaboration with your student research partner been refunded/offered? |
| Please describe any aspects of the Student Patient Alliance that could be improved: |
| Please describe any additional thoughts about your collaboration with your student research partner that have not been captured in this form: |

Supplementary Materials 2: Record PPI activities

### **<u>Student Patient Alliance – activity log</u>**

Please complete the following table to record in detail your patient/public involvement activities. Insert additional rows whenever necessary.

| **Student name:** | | **University attended:** |
|---|---|---|

| **Purpose of activity** (E.g. develop lay summary/news article, discuss research priorities…) | **Type of activity** (E.g. face to face meeting; email exchange…) | **Who initiated the activity?** (E.g. student, patient partner, supervisor, PPI lead…) | **Date(s) & time of activity** (Please include the time spent to the nearest 30 mins) | **Impact of activity** (E.g. changes made to research design/documents; things you have learnt, skills you have developed as a result of the interaction) | **Date(s) feedback on impact of activity given to patient partner** | **Type of feedback** (E.g. email message, phone call) | **Comments** (E.g. challenges encountered, solutions implemented, training needs identified, future plans…) |
|---|---|---|---|---|---|---|---|

Supplementary Materials 3: Web-based feedback forms for students and their PPI partners.

**Student Patient Alliance**

**Feedback form for students (hard copy of the Microsoft Forms questions)**

Please complete the following form every 6 months to feedback on your patient/public involvement.

Your form will be shared with specific members of staff at your affiliated university. Your answers will remain anonymous. Please return to Rebecca Birch

| | |
|---|---|
| **Date form completed** | |
| **Name** | |
| **Which university do you attend?** | ☐University of Birmingham<br>☐University of Glasgow<br>☐Newcastle University<br>☐University of Nottingham<br>☐University of Oxford<br>☐University of Southampton<br>☐ Other (please specify) |
| **Year of study** | ☐Year 1<br>☐Year 2<br>☐Year 3<br>☐Year 4<br>☐ Other (please specify) |
| **Which of the following best describes your PhD research?** | ☐Laboratory based research<br>☐Experimental medicine research<br>☐Clinical research<br>☐Health services research<br>☐ Other (please specify) |
| **Start date of your PhD project** | DD/MM/YYY |
| **End date of your PhD project** | DD/MM/YYY |

| **Training and support** | |
|---|---|
| **Which of the following resources did you have access to? Tick all that apply** | ☐R2P2 and Student Patient/Public Alliance resources<br>☐Research funder resources (e.g. Versus Arthritis)<br>☐INVOLVE / NIHR resources<br>☐I didn’t receive any resources<br>☐ Other (please specify) |
| **How useful were the resources you received?** | ☐Very useful<br>☐Useful<br>☐Neutral<br>☐Not useful |

| | |
|---|---|
| | ☐Not at all useful<br>☐Not applicable - I didn't receive any resources |
| **Please describe any additional resources/information that would have been useful** | |
| **Please describe any additional training about patient/public involvement in research that you would like to receive** | |
| **What administrative support have you received? Tick all that apply** | ☐Support with voucher / bank transfer payments to patient/public partners<br>☐Support with reimbursement of patient/public partner expenses<br>☐Arranging face to face meetings with patient/public partners<br>☐ Arranging telephone or video calls with patient/public partners<br>☐Providing information about patient/public involvement and training opportunities<br>☐None<br>☐Other (please specify) |

| **Impact** | | | | | | |
|---|---|---|---|---|---|---|
| **How much do you agree with the following statements?<br>Taking part in the Student Patient/Public Alliance has:** | | | | | | |
| | **Strongly agree** | **Agree** | **Neutral** | **Disagree** | **Strongly disagree** | **N/A<br>I have not interacted with my patient/public partner(s)** |
| Improved my knowledge about patient/public involvement in research | ☐ | ☐ | ☐ | ☐ | ☐ | ☐ |
| Improved my confidence to communicate about my research with members of the public | ☐ | ☐ | ☐ | ☐ | ☐ | ☐ |

| | | |
|---|---|---|
| Changed aspects of my research design | | |
| Increased my interest in my research area | | |
| Shaped my future research/career plans | | |
| Improved my understanding of research ethics | | |
| Improved my knowledge of patient/public priorities for research | | |
| **To what extent has your collaboration with your patient/public research partner(s) had a positive/negative impact on you / your research overall?** | | ☐Very positive impact<br>☐Positive impact<br>☐Neither positive nor negative impact<br>☐Negative impact<br>☐Very negative impact |
| **Please describe any positive impact on you/your research overall** | | |
| **Please describe any negative impact on you/your research overall** | | |
| **What was the background of your patient/public partner(s)?** | | ☐ My partner(s) was a patient with a disease relevant to my research area<br>☐ My partner(s) was a member of the public. Any medical conditions they had were not relevant to my research area |

| **Patient partners** | |
|---|---|
| **How much do you agree with the following statement: Taking part in the Student Patient/Public Alliance has improved my understanding of my partner's disease** | ☐ Strongly agree<br>☐ Agree<br>☐ Neutral<br>☐ Disagree<br>☐ Strongly disagree<br>☐ I haven't interacted with my patient/public research partner |

| | |
|---|---|
| | ☐ Not applicable – my partner was not a patient with a disease relevant to my research |
| **Public partners** | |
| **How much do you agree with the following statement: Taking part in the Student Patient/Public Alliance has improved my understanding of my research from the public's point of view** | ☐ Strongly agree<br>☐ Agree<br>☐ Neutral<br>☐ Disagree<br>☐ Strongly disagree<br>☐ I haven't interacted with my patient/public research partner |

| | |
|---|---|
| **Concluding thoughts** | |
| **How did you hear about the Student Patient/Public Alliance?** | ☐ My supervisor<br>☐ My centre manager / other university staff<br>☐ Word of mouth<br>☐ Other (please specify) |
| **How likely are you to recommend the Student Patient/Public Alliance to other research students?** | ☐ Very likely<br>☐ Likely<br>☐ Neither likely nor unlikely<br>☐ Not likely<br>☐ Not very likely |
| **Please describe any aspects of the Student Patient/Public Alliance that worked well** | |
| **Please suggest aspects of the Student Patient/Public Alliance that could be improved** | |
| **Please describe any additional thoughts about the Student Patient/Public Alliance that have not been captured in this form** | |

**Student Patient Alliance**

**Feedback form for patients / members of the public (hard copy of the Microsoft Forms questions)**

Please complete the following form every 6 months to feedback on your patient/public involvement.

Your form will be shared with specific members of staff at your affiliated university. Your answers will be collated and anonymised. Please return to Rebecca Birch

| | |
|---|---|
| **Date form completed** | |
| **Name** | |
| **Which university does your PhD student attend?** | ☐University of Birmingham<br>☐University of Glasgow<br>☐Newcastle University<br>☐University of Nottingham<br>☐University of Oxford<br>☐University of Southampton<br>☐ Other (please specify) |
| **Please indicate which (if any) of the following research activities you have been involved in with your student partner(s). Tick all that apply.** | ☐ Developing plain English summaries of research<br>☐ Developing grant applications<br>☐ Developing research posters<br>☐ Developing ethics applications<br>☐ Developing research presentations<br>☐ Contributing to scientific publications<br>☐ Contributing to events about research<br>☐ Advising on research design / analysis / findings<br>☐ Developing research priorities / ideas<br>☐ Advising on research documents<br>☐ Other (please describe):________________________________ |
| **Please select a number to indicate how satisfied you are with the amount of interaction that you have had with your student partner** | 1 2 3 4 5 6 7 8 9 10<br><br>1 – not at all satisfied<br>10 – extremely satisfied |
| **Please describe what went well:** | |

| | |
|---|---|
| **Please describe what could be improved:** | |
| **How much do you agree with the following statement: I have received feedback from my student partner(s) on the impact of my involvement in their project(s).** | ☐ Strongly agree<br>☐ Agree<br>☐ Neutral<br>☐ Disagree<br>☐ Strongly disagree |
| **In what ways has your collaboration with your student partner(s) had a positive or negative impact on you overall?** | ☐ Very positive impact<br>☐ Positive impact<br>☐ Neither positive nor negative impact<br>☐ Negative impact<br>☐ Very negative impact |
| **Please describe any positive impact** | |
| **Please describe any negative impact** | |
| **Please record any training / learning opportunities about patient/public involvement in research that you have undertaken/received:** | ☐ Student Patient / Public Alliance information materials<br>☐ Courses / workshops about patient / public involvement in research<br>☐ Other (please describe):________________________________ |
| **Please describe any additional training about patient/public involvement in research that you would like to receive:** | |
| **Have you incurred any expenses as a result of your collaboration** | ☐ Yes<br>☐ No<br>☐ Don’t know |

| | |
|---|---|
| **with your student research partner?** | |
| **Have any expenses you have incurred been refunded/offered?** | ☐ Yes<br>☐ No<br>☐ Don't know |
| **How did you hear about the Student Patient Alliance?** | ☐ Word of mouth<br>☐ Local patient / public involvement group<br>☐ Promotional email or website<br>☐ Social media<br>☐ Other (please describe):________________________________ |
| **How likely are you to recommend the Student Patient Alliance to other patients / members of the public?** | ☐ Very likely<br>☐ Likely<br>☐ Neither likely nor unlikely<br>☐ Not likely<br>☐ Not very likely |
| **Please describe any aspects of the Student Patient Alliance that worked well** | |
| **Please describe any aspects of the Student Patient Alliance that could be improved:** | |
| **Please describe any additional thoughts about your collaboration with your student research partner that have not been captured in this form:** | |

Supplementary materials 4. Quotes pilot SPA evaluation

| | |
|---|---|
| **Specific opportunities for collaboration helpful for partnership** | |
| **1** | *"It was quite good to have something to work towards, to work on together and it was just a summary of my research and so it was good for me because it made me understand it better to explain it. But also, it was a good process, a tool for talking to other people"* (Student) |
| **Mutual learning and support** | |
| **1** | *"With you having to explain it all from the basic, absolute basic gave me a greater understanding… when you teach you learn don't you? It's a win-win really".* (PRP) |
| **2** | *"It kept my focus on the bench to bedside aspect because I think us being basic science researchers, we're really focused on getting the right research design, having the right experiments, testing everything at a very molecular level and I think we tend to forget that the only reason that we're doing this is to actually improve patients' lives… And it does bring my motivation up quite a bit, particularly if things aren't going terribly well in the labs or if I'm in a transition part of the project."* (Student) |
| **3** | *"I could see, yes you were getting somewhere with it and there's almost a feeling of excitement that when we next meet up in the next couple of months or so - I'm interested to see, has that line of research gone further. And maybe with everybody that's studying and working its quite good to have perhaps some positive feedback and I think the patient partners can do that. Because you do feel this gratitude for the fact that somebody's doing something that might help you but probably more likely help other people."* (PRP) |
| **4** | *"There are a lot of days you have not good days in the lab, things aren't going well, but when you have these meetings, you can see like it is having a positive effect, or you get positive feedback, or someone's interested in your research and asking questions on this, that, everything. It makes you think, but it also makes you realise that what you're doing is good."* (Student) |

| Considerations for future SPA initiatives | |
|---|---|
| **1** | *"There might be complications, but probably you know students wouldn't be getting in touch, or not collaborating… If it comes from the funder – top down – everyone has to do it."* (Student) |
| **2** | *"Just being able to organise our own meetings face-to-face when suits us best, and the location that suits us best because there's no specific structure, it just means we can have a chat, and then we can move on to the research. I think that's worked really well, it just means you can form that sort of good relationship".* (Student) |
| **3** | *"It would probably be good if, like when you're setting up partners if there's a question of: Ok you do animal work, and if they're ok with that, just in case… if someone didn't like animal research and didn't want to discuss it, if your research is all on that, and you haven't had that discussion."* (Student) |
